# Context-specific eQTLs reveal causal genes underlying shared genetic architecture of critically ill COVID-19 and idiopathic pulmonary fibrosis

**DOI:** 10.1101/2024.07.13.24310305

**Authors:** Trisha Dalapati, Liuyang Wang, Angela G. Jones, Jonathan Cardwell, Iain R. Konigsberg, Yohan Bossé, Don D. Sin, Wim Timens, Ke Hao, Ivana Yang, Dennis C. Ko

**Author notes:** To whom correspondence should be addressed: Dennis C. Ko, 0048B CARL Building Box 3053, 213 Research Drive, Durham, NC 27710. @denniskoHiHOST.

## Abstract

Most genetic variants identified through genome-wide association studies (GWAS) are suspected to be regulatory in nature, but only a small fraction colocalize with expression quantitative trait loci (eQTLs, variants associated with expression of a gene). Therefore, it is hypothesized but largely untested that integration of disease GWAS with context-specific eQTLs will reveal the underlying genes driving disease associations. We used colocalization and transcriptomic analyses to identify shared genetic variants and likely causal genes associated with critically ill COVID-19 and idiopathic pulmonary fibrosis. We first identified five genome-wide significant variants associated with both diseases. Four of the variants did not demonstrate clear colocalization between GWAS and healthy lung eQTL signals. Instead, two of the four variants colocalized only in cell-type and disease-specific eQTL datasets. These analyses pointed to higher *ATP11A* expression from the C allele of rs12585036, in monocytes and in lung tissue from primarily smokers, which increased risk of IPF and decreased risk of critically ill COVID-19. We also found lower *DPP9* expression (and higher methylation at a specific CpG) from the G allele of rs12610495, acting in fibroblasts and in IPF lungs, and increased risk of IPF and critically ill COVID-19. We further found differential expression of the identified causal genes in diseased lungs when compared to non-diseased lungs, specifically in epithelial and immune cell types. These findings highlight the power of integrating GWAS, context-specific eQTLs, and transcriptomics of diseased tissue to harness human genetic variation to identify causal genes and where they function during multiple diseases.

## INTRODUCTION

Genome-wide association studies (GWAS) are a powerful approach to identify relationships between disease phenotypes and genetic variants, most commonly single nucleotide polymorphisms (SNPs). Elucidating associated SNPs can reveal unanticipated underlying pathophysiology of disease, unbiased by preconceived notions of plausible biology, and may lead to informed drug targets and therapies. Indeed, success along the drug development pipeline increases with human genetic evidence.^1^ In 2021, two-thirds of the Food and Drug Administration approved drugs were supported by human genetic evidence.^2^ Beyond associations with individual diseases, multiple independent GWAS can be integrated to identify pleiotropy – the phenomenon where a single genetic locus affects multiple diseases or traits that are otherwise considered unrelated.^3,4^ Pleiotropy can reveal shared pathogenic mechanisms among different diseases. Understanding these mechanisms and identifying potential therapeutic targets require the identification of the causal genes associated with the genetic variant.

One method to identify putative casual genes is to measure the degree of colocalization between disease GWAS and molecular quantitative trait loci (QTLs), such as expression QTLs (eQTLs), using statistical frameworks that integrate summary statistics to determine if overlapping signals are due to the same causal SNP. ^5-7^ While the chain of causality implied by colocalization makes intuitive sense (a SNP affects transcription of a gene to impact disease), there is surprisingly poor colocalization between disease GWAS and eQTL signals.^8^ One possible reason for lack of colocalization includes using eQTL datasets from unsuitable contexts. For example, cell- and disease-specific eQTLs may be required.^8^ Alternatively, other kinds of molecular QTLs, such as methylation, splicing, or protein QTLs (mQTLs, sQTLs, pQTLs, respectively), may be more causally important in certain disease pathogenesis.^8,9^

Here, we focused on pleiotropic SNPs associated with both critically ill COVID-19 and idiopathic pulmonary fibrosis (IPF) and formally tested colocalization with context-specific eQTLs to identify the likely causal genes. We previously developed a software called the interactive Cross-Phenotype Analysis of GWAS database (iCPAGdb) to study pleiotropy by comprehensively identifying shared association signals between user-uploaded GWAS and all publicly cataloged GWAS summary statistics.^10,11^ Using the first COVID-19 GWAS published by Ellinghaus et al.^12^ and the highest-powered IPF GWAS at the time,^13^ we identified a shared signal in *DPP9*, rs12610495, associated at a suggestive threshold with severe COVID-19 (p-value = 5.20 x 10^-6^)^12^ and at genome-wide significance with IPF (p = 2.92 × 10^−12^)^13^.^11^ The *DPP9* locus was later confirmed to be associated with critically ill COVID-19 at genome-wide significance.^14-16^ Others have also confirmed and expanded on the shared genetic associations between critically ill COVID-19 and IPF.^11,17-19^

Notably, post-COVID-19 pulmonary fibrosis (PCPF) is clinically reminiscent of IPF.^20^ PCPF, which occurs due to irreversible lung scarring and stiffening, causes progressive difficulties in breathing and ultimately necessitates lung transplantation.^20,21^ Critical illness is a major risk factor for PCPF.^22^ A meta-analysis reported that nearly 45 percent of COVID-19 survivors had long-term respiratory symptoms and fibrosis, emphasizing the urgent need to understand the mechanisms and potential treatments of PCPF.^22^ In fact, drugs for IPF, including pirfenidone and nintedanib, have been co-opted during emergency treatment of PCPF.^23^ Thus, shared genetic associations between critically ill COVID-19 and IPF might reflect common pathophysiology that could be utilized for therapeutic benefit in both diseases.

In this study, we leveraged iCPAGdb again with the most current, highest-powered COVID-19 GWAS (COVID-19 HGI release 7)^24^ and IPF meta-GWAS^25^ to determine whether additional loci are associated with both diseases. We have identified five shared loci, including the *DPP9* locus containing rs12610495, that are likely due to the same causal variants based on colocalization analysis. For two of the variants, their risk alleles are reversed in COVID-19 and IPF, highlighting that a shared signal may have different functions in the two diseases and that the pulmonary damage from COVID-19 is likely distinct from IPF. We systematically identified the likely causal genes underlying the shared genetic architecture between critically ill COVID-19 and IPF by performing colocalization analysis using bulk, single-cell, and disease-specific eQTL datasets. For two genes, *ATP11A* and *DPP9,* we found colocalization of GWAS and eQTL signals only in a cell-type and disease-specific context. Therefore, context-specific eQTL are critical for identifying the causal genes underlying the shared genetic architecture of critically ill COVID-19 and IPF and may lead to new insights connecting the diseases.

## MATERIALS AND METHODS

### GWAS summary statistics and QTL datasets

All datasets used in this study and accession information are summarized in **Table 1** and **Table S1**. For analyses using GTEx v8, we obtained the “Tissue-Specific All SNP Gene Associations” files via the Google Cloud Platform, which included non-significant associations not searchable on the web browser.^26,27^ In addition to the IPF eQTL and mQTL summary statistics, Borie et al. provided *DPP9* expression and cg07317664 methylation data.^28^ The genome coordinates in datasets were lifted to GRCh38 using the R packages “liftOver”^29^ and “rtracklayer”^30^.

**Table 1:** Datasets used for present study.

| Dataset | Article Author (Year) | Type |
| --- | --- | --- |
| Very severe respiratory confirmed (critically ill) COVID-19 vs. population | COVID-19 Host Genetics Initiative (2021) | GWAS |
| 5-way meta-GWAS of IPF susceptibility | Allen (2022) | GWAS |
| GTEx Conditionally Independent eQTL | Lonsdale (2013); GTEx Consortium (2017, 2020) | eQTL |
| GTEx Tissue-Specific All SNP Gene Associations eQTLs | Lonsdale (2013); GTEx Consortium (2017, 2020) | eQTL |
| Human Lung Tissue eQTL Study | Hao (2012) | eQTL |
| BLUEPRINT | Chen (2016) | eQTL |
| Bulk IPF | Borie (2022) | eQTL; mQTL |
| Single-cell COVID-19 PBMCs | Aquino (2023) | eQTL |
| Single-cell IPF Lung | Natri (2023) | eQTL |
| IPF Lung Study 1 | Jaffar (2022) | Bulk RNA-Seq |
| IPF Lung Study 2 | Ammar (2019) | Bulk RNA-Seq |
| COVID-19 Whole Blood Study | Li (2021) | Bulk RNA-Seq |
| COVID-19 Lung Study | de Rooij (2022) | Bulk RNA-Seq |
| IPF Cell Atlas | Adams (2020) | scRNA-Seq |
| Lung Atlas of Lethal COVID-19 | Melms (2021) | snRNA-Seq |
| Abbreviations: GTEx, Genotype-Expression Project; SNP, single nucleotide polymorphism; GWAS, genome-wide association studies; eQTL, expression quantitative trait loci; mQTL, methylation quantitative trait loci; RNA-seq, RNA-sequencing; scRNA, single-cell RNA; snRNA, single-nucleus RNA |  |  |

### Discovery of pleiotropic SNPs using iCPAGdb

We previously reported the development of iCPAGdb which identified cross-phenotypic associations across 4400 traits loaded from the NHGRI-EBI GWAS Catalog.^10,11^ For this study, critically ill COVID-19 summary statistics were formatted for the web browser (http://cpag.oit.duke.edu/explore/app/) and uploaded as the User Supplied GWAS for “GWAS source one”. NHGRI was selected for “GWAS source two”. P-threshold_1_ (factor_1_ X 10-^x^_1_) and P-threshold_2_ (factor_2_ X 10-^x^_2_) were set to genome-wide significant (5x10^-8^; 5 for “factor_1_” and “factor _2_” and 8 for “x_1_” and “x_2_”). “European” was chosen for “LD 1000 Genomes population” as the linkage disequilibrium (LD) population as the IPF GWAS was based on European ancestry. Additionally, the command line version of iCPAGdb (https://github.com/tbalmat/iCPAGdb) was utilized to directly compare the critically ill COVID-19 and IPF meta-GWAS, which became publicly available after the publication of iCPAGdb. The full results from iCPAGdb after uploading the critically ill COVID-19 summary statistics are listed in **Table S2**.

### Identification of candidate eGenes and LD proxies

The five SNPs shared between COVID-19 and IPF identified by iCPAGdb were queried in the eQTL datasets in **Table 1**. We checked whether each SNP was a conditionally independent eQTL in each of the 54 tissues in GTEx v8. Next, using the GTEx web browser, we downloaded all eGenes associated with each SNP. We then searched for conditionally independent eQTLs across the 54 tissues associated with the list of eGenes. We used LDmatrix to check if any of the conditionally independent eQTLs were in LD with our SNPs and could serve as LD proxies (r^2^ > 0.50) (**Table S3**).

For rs12585036 and rs12610495, the only conditionally independent SNPs in LD were eQTLs for *ATP11A* and *DPP9*, respectively. Two conditionally independent SNPs were in perfect LD with rs2897075 and were eQTLs for *ZKSCAN1* and *COPS6*. rs1105569 was in strong LD (r^2^ > 0.90) with 330 conditionally independent eQTL associated with 23 eGenes across 49 tissues, underscoring the challenges in identifying causal genes in the 17q21.31 inversion supergene.^31^ For this locus, we further filtered conditionally independent eQTL in strong LD in tissues of interest for COVID-19 and IPF pathogenesis– fibroblasts, lung, lymphocytes, and whole blood. From literature review, we identified one additional eGene, *TRIM4*, associated with rs2897075^17^ and two additional eGenes, *CRHR1* and *SPPL2C*, associated with rs1105569^31^. Altogether, we focused on 15 candidate protein-coding eGenes for our five SNPs of interest **(Table 2)**.

**Table 2:**
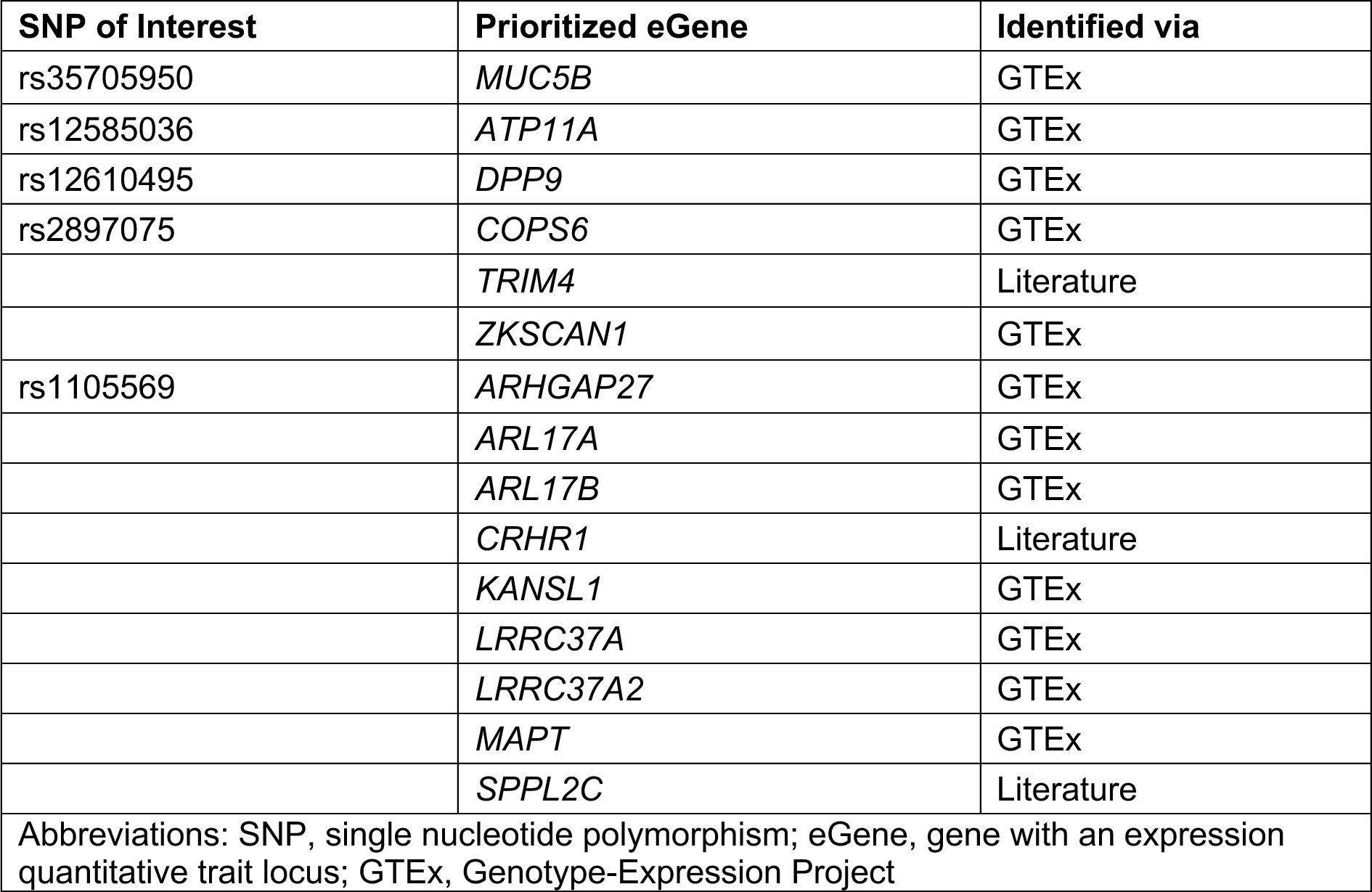
Candidate protein-coding eGenes associated with SNPs of interest.

### Colocalization analysis

We applied Giambartolomei et al.’s colocalization analysis (COLOC), using the R package “coloc”,^6^ to determine if the associations identified by iCPAGdb were due to the same causal SNP. COLOC uses a Bayesian framework to calculate the posterior probabilities that two traits are not associated in the locus of interest (PP0), only one trait is associated in the locus (PP1 and PP2), both traits are associated at the locus but with different, independent causal variants (PP3), or both traits are associated with a single causal variant in the locus (PP4). For COLOC using only GWAS summary statistics, we filtered SNPs within a 1 megabase (Mb) window from the SNP of interest. For COLOC using eQTL datasets, we filtered all eQTLs for a candidate eGene and then filtered SNPs within a 1 Mb window from the SNP of interest. We ran the COLOC “coloc.abf” function using the default prior parameters, p1 = 1 × 10^-4^, p2 = 1 × 10^-4^, and p12 = 1 × 10^-5^ for all analyses. PP4 between 0.700 to 0.900 was interpreted as likely to share a single causal variant, while PP4 > 0.900 was interpreted as sharing a single causal variant. The PP4/PP3 measured the intensity of the colocalization signal with values ≥5.00 indicating further support for colocalization and ≥3.00 suggesting likely colocalization.^9,32^

### IPF and COVID-19 transcriptomic analysis

We utilized publicly available bulk RNA datasets from lung or blood from patients diagnosed with COVID-19 and IPF. Normalized counts were downloaded from GEO (GSE213001, GSE134692, GSE172114).^33-35^ When normalized counts were unavailable, raw counts were downloaded (GSE159585).^36^ Genes with counts less than five were filtered out, and the remaining gene counts were normalized using DESeq2^37^. We utilized publicly available single-cell (sc) and single-nucleus (sn) RNA-sequencing (RNA-Seq) datasets to investigate causal gene expression in IPF and COVID-19 lungs compared to healthy controls. A scRNA-Seq dataset for IPF from Adams et al.^38^ containing 243,472 cells from 32 IPF lungs and 28 control donor lungs was downloaded from GEO (GSE136831). A snRNA-Seq dataset for lethal COVID-19 from Melms et al.^39^ containing 116,314 nuclei from 19 lethal COVID-19 lungs and seven control lungs was downloaded from the Broad Institute Single Cell Portal.^40^ Previously annotated cell type clusters were used to create expression dot plots and calculate cell type proportions within each donor with the Python package “scanpy”.^41^ Only cell type clusters with at least ten cells and cells with at least 500 total counts were retained for downstream differential expression analyses. We then pseudobulked gene counts by donor and cell-type using the Python package “decoupleR”^42^. Differential expression analysis between diseased and healthy lungs was conducted in the Python package “pyDESeq2”^43^ which implements the traditional DESeq2^37^ workflow. Cell type proportions calculated from the datasets are in **Table S4-5**. Differential expression results are in **Table S6-7**.

### Data Visualization

All visualizations were created in R version 4.3.0.^44^ Lollipop plots, genotypic box plots and -log_10_(p-value) scatterplots were generated using “ggplot2”.^45^ Manhattan plots and LocusZoom plots were created using “fastman”^46^ and “locuszoomr”^47^, respectively. While p-values from eQTL and mQTL summary statistics incorporated PEER factor normalization, values plotted for gene expression and methylation were pre-PEER normalization. For eQTL data from Borie et al.,^28^ normalized RNA TPM values were plotted (transcripts per million after trimmed mean of M values (TMM) normalization across samples and inverse normal transformation on a per-gene basis). For mQTL data from Borie et al.,^28^ normalized methylation beta referred to beta values of DNA methylation level measured on Illumina arrays (scale 0–1) after SeSame^48^ data preprocessing and normalization.

## RESULTS

### Identification of loci shared between critically ill COVID-19 and idiopathic pulmonary fibrosis

The underlying causal genes and mechanisms for the shared loci associated with critically ill COVID-19 and IPF have not been systematically identified. The COVID-19 Human Genetics Initiative integrated 82 studies from 35 countries to create the largest cohort to date for risk, hospitalization, and critical illness.^24,49,50^ Herein, we studied the critically ill COVID-19 trait (cases = 18,152, controls = 1,145,546) as severe disease is a risk factor for developing lung fibrosis. iCPAGdb revealed shared genetic associations between critically ill COVID-19 and multiple human diseases (**Figure 1A; Table S2**). Most interestingly, critically ill COVID-19 significantly overlapped with several pulmonary traits (**Figure 1B)**, including interstitial lung disease (ILD) and IPF at five loci (ILD: Fisher’s exact test FDR = 6.05 x 10^-15^ and 3774.0-fold enrichment; IPF: FDR = 1.56 x 10^-14^ and 4780.0-fold enrichment) (**Figure 1C-D**). Importantly, the risk allele was reversed for two of the five SNPs, rs35705950 and rs12585036. The odds ratio at each shared locus was also generally larger for IPF, highlighting the complex disease mechanisms linking COVID-19 and IPF (**Figure 1C**).

**Figure 1:**
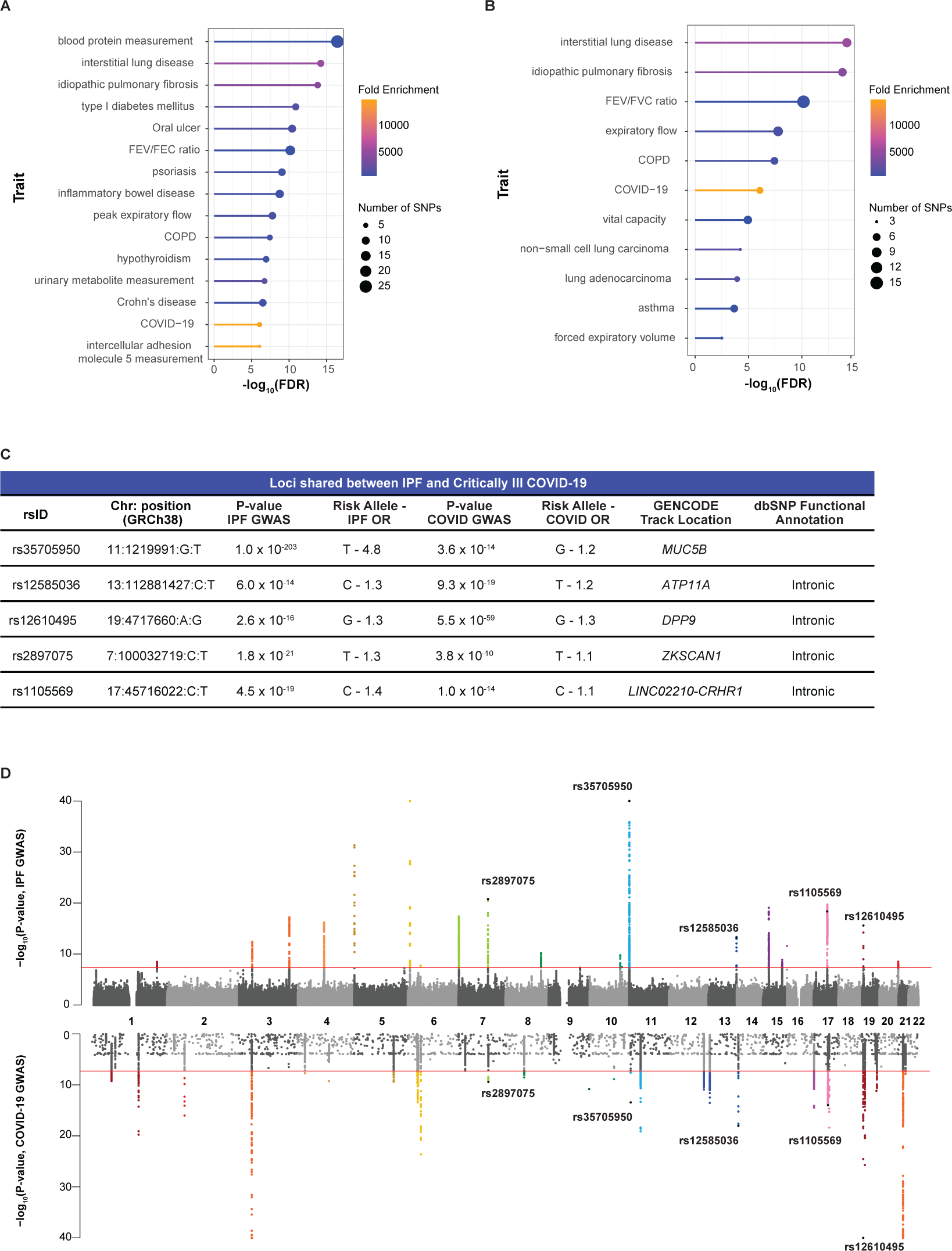
iCPAGdb reveals associations with critically ill COVID-19. (A) Lollipop plot of the top 15 traits associated with COVID-19. Fold enrichment is calculated by dividing number of SNPs shared by number of SNPs expected to be shared. (B) Plot of the pulmonary traits associated with COVID-19. (C) Table of the five SNPs overlapping between critically ill COVID-19 and IPF. The location and functional annotation are from GENCODE on the UCSC Genome Browser and NCBI dbSNP, respectively. (D) Miami plot of the IPF and critically ill COVID-19 GWAS highlights the five shared SNPs. Over 7.5 million SNPs were included in the IPF meta-GWAS summary statistics, while the top 10,000 SNPs were publicly available in the critically ill COVID-19 summary statistics from the COVID-19 Human Genetics Initiative. P-values are -log_10_ transformed, and the red line indicates the genome-significant threshold at p < 5.0 × 10^-8^.

Although regional association plots visually suggested colocalizing signals in critically ill COVID-19 and IPF (**Figure 2A-E**), shared associations cannot be assumed to be driven by the same causal SNP. Overlapping GWAS signals may be due to a shared causal SNP or two independent signals driven by different causal SNPs with variable degrees of LD. To systematically reveal whether each signal was due to a single causal SNP, we performed formal colocalization testing using the critically ill COVID-19 GWAS and the IPF meta-GWAS (cases = 4,125, controls = 20,464). For all loci except rs1105569, the PP4 was greater than 0.900 (**Figure 2F; Table S9**). The exception, rs1105569 (PP4 = 0.725), is located within the 17q21.31 inversion supergene with extensive LD extending over nearly 900 kb,^31^ which precluded our ability to determine whether an individual causal variant was driving the association in both diseases (**Figure 2E).** Overall, we determined that for the five shared loci identified by iCPAGdb, four demonstrated colocalizing signals likely driven by the same causal SNP in both critically ill COVID-19 and IPF.

**Figure 2:**
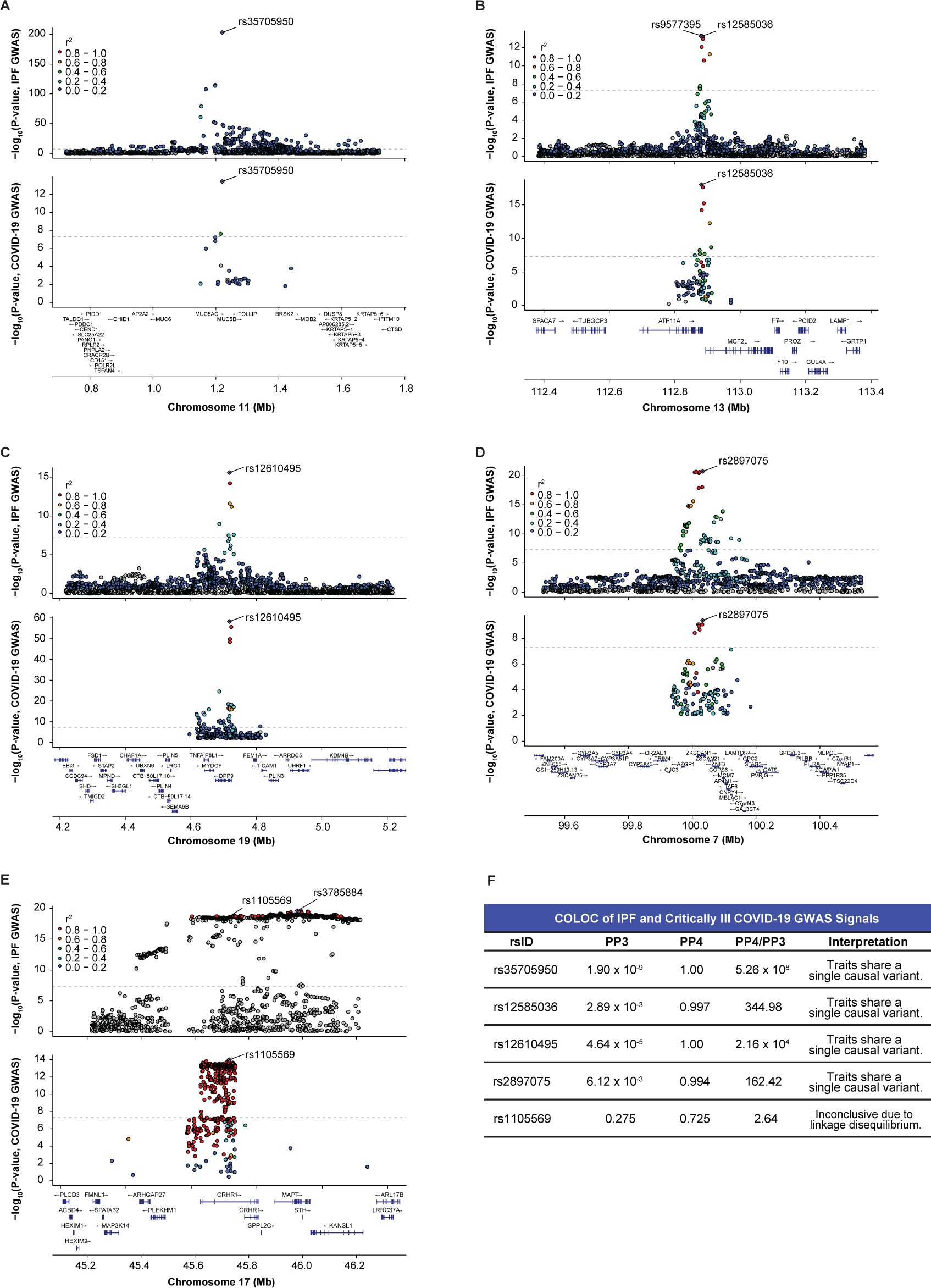
IPF and critically ill COVID-19 GWAS colocalize at shared signals. (A) LocusZoom plot highlights that rs35705950 is the lead variant (purple diamond) in both disease GWAS and is located upstream of *MUC5B* on chromosome 11. (B) rs12585036 is a top variant in the IPF GWAS and is in strong LD with the lead variant (rs9577395). rs12585036 is the lead variant in the COVID-19 GWAS and is located within an intron of *ATP11A* on chromosome 13. (C) rs12610495 is the lead variant in both disease GWAS and is located within an intron of *DPP9* on chromosome 19. (D) rs2897075 is the lead variant in both disease GWAS and is located within an intron of *ZKSCAN1* on chromosome 7. (E) rs1105569 is a top variant in the IPF GWAS and is in strong LD with the lead variant (rs3785884). rs1105569 is the lead variant in the COVID-19 GWAS. rs1105569 is located within an intron of *CRHR1* and in a supergene with high linkage disequilibrium between SNPs. (F) COLOC indicates strong colocalization between the disease GWAS signals at four of the five shared SNPs with PP4 > 0.900, and PP4/PP3 > 5.00. COLOC is inconclusive for rs1105569 due to extensive linkage disequilibrium in the region. P-values are -log_10_ transformed, and the dotted line indicates the genome-significant threshold at p < 5.0 × 10^-8^. Linkage disequilibrium information for European populations was obtained from LDlink and is relative to the lead variant in each plot.

### Identification of causal genes for shared loci in bulk tissue

SNPs can affect a nearby gene through altering function or regulation. To speculate how each SNP contributed to gene regulation during COVID-19 and IPF, we used HaploReg,^51^ a publicly available tool that annotates the putative functional impact of queried SNPs and all SNPs in LD. We first explored whether any of our SNPs of interest were in strong LD with nonsynonymous variants, altering the amino acid sequence of the protein. For rs1105569, the lead variant was in strong LD (r^2^>0.80) with 18 nonsynonymous variants (missense, frameshift, and nonsense) in *CRHR1*, *SPPL2C*, and *MAPT*.^51^ These three genes, and several others in 17q21.31, were previously considered important for COVID-19.^14,24,50^ No nonsynonymous variants were in strong LD with the other four SNPs of interest, indicating these signals were likely due to regulatory variants.

Next, we determined if the five SNPs of interest were associated with gene expression in bulk, healthy tissue. In GTEx,^26^ only rs35705950 was a conditionally independent eQTL for *MUC5B* in lung (nominal p-value = 6.71 x 10^-16^). We next assessed if our SNPs of interest were in LD with conditionally independent eQTLs in any tissue (**Table S2**). Through GTEx and literature review, we identified 15 plausible eQTL-protein-coding eGene pairs for COLOC analysis (**Table 2**).

A previous study informed that rs35705950 is within an enhancer region of *MUC5B* that was differentially methylated and bound by the transcription factor FOXA2.^52^ Subsequent colocalization analyses with eQTL and mQTL data from control and IPF lung tissue revealed that the T allele of rs35705950 was also associated with higher methylation within a repressor region of *MUC5B* and higher *MUC5B* expression.^28^ Similarly, we found that *MUC5B* was the only eGene for rs35705950 in the lung. Colocalization of the disease GWAS and *MUC5B-*eQTL signals resulted in a PP4 of 1.00, indicating that rs35705950 was the causal SNP driving both the GWAS and eQTL signals (**Figure 3A-B**).

**Figure 3:**
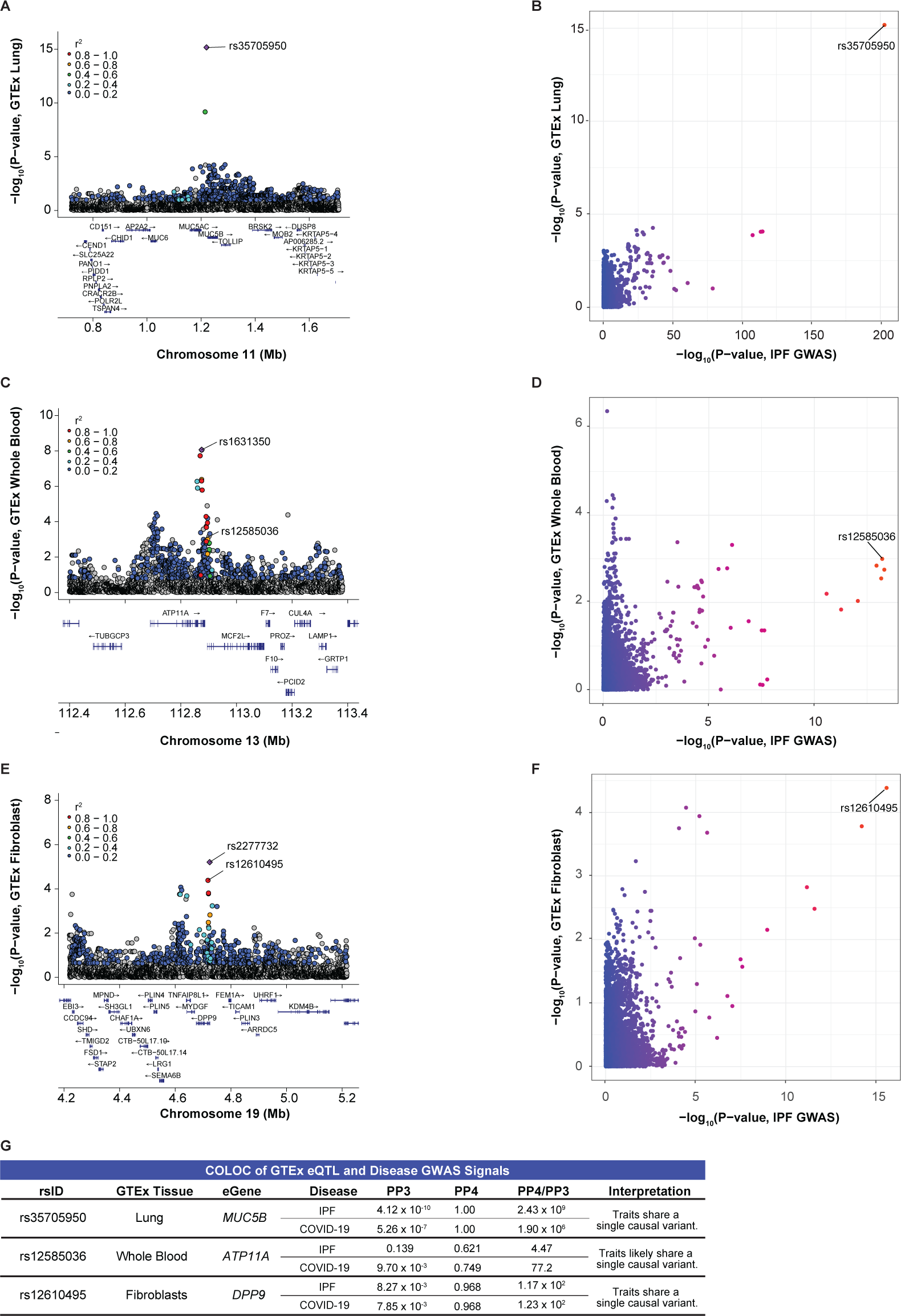
Colocalization of Disease GWAS and GTEx eQTL signals. (A) LocusZoom plot shows rs35705950 is the lead eQTL (purple diamond) for *MUC5B* in GTEx lung tissue. (B) Comparison of -log_10_(p-values) from the IPF GWAS and *MUC5B-*eQTLs in GTEx lung tissue shows rs35705950 as the shared lead SNP. (C) rs12585036 is not the lead eQTL nor in LD with the lead eQTL (rs1631350) for *ATP11A* in GTEx whole blood. (D) Comparison of -log_10_(p-values) from the IPF GWAS and *ATP11A-*eQTLs in GTEx lung tissue shows rs12585036 as the shared lead SNP. (E) rs12610495 is in strong LD with the lead eQTL (rs2277732) for *DPP9* in GTEx fibroblasts. (F) Comparison of -log_10_(p-values) from the IPF GWAS and *DPP9-*eQTLs in GTEx fibroblasts show rs12610495 as the shared lead SNP. (G) COLOC indicates strong colocalization between the disease GWAS and *MUC5B- and DPP9-*eQTL signals in lung and fibroblasts, respectively, with PP4 > 0.900 and PP4/PP3 > 5.00. COLOC suggests colocalization between the COVID-19 GWAS and *ATP11A*-eQTL signals in whole blood with PP4 > 0.700 and PP4/PP3 > 5.00. P-values are -log_10_ transformed. Linkage disequilibrium information for European populations was obtained from LDlink and is relative to the lead variant in each plot.

An LD proxy of rs12585036, rs9577395 (r^2^=0.99) was a conditionally independent eQTL for *ATP11A* in tissues of unclear relevance to IPF and COVID-19 (aorta, skin, small intestine). However, in tissues relevant to IPF and COVID-19 (EBV-transformed lymphocytes, cultured fibroblasts, lung, and whole blood), we found modest colocalization of the GWAS and *ATP11A-*eQTL signals in whole blood (PP4 = 0.621 with IPF GWAS; 0.749 with COVID-19 GWAS; **Figure 3C-D)**. In the remaining three tissues, we did not find colocalization of the GWAS and *ATP11A-*eQTL signals (PP4 < 0.408; **Table S8**), and the lead eQTLs were in weak LD with rs12585036 (r^2^ < 0.20).

An LD proxy of rs12610495, rs2277732 (r^2^ = 0.95) was a conditionally independent eQTL for *DPP9* in fibroblasts. Accordingly, the disease GWAS and *DPP9-* eQTL signals colocalized in fibroblasts (PP4 = 0.989), highlighting that this variant may affect *DPP9* expression in a cell-type specific manner (**Figure 3E-F**). In GTEx, the G allele was associated with lower *DPP9* expression in lungs and fibroblasts and was also the risk allele for both critically ill COVID-19 and IPF. In the remaining three tissues, we did not find colocalization of the GWAS and eQTL signals (PP4 < 0.225; **Table S8**), and the lead eQTLs were in weak LD with rs12610495 (r^2^ < 0.20).

LD proxies of rs2897075, rs73158411 and rs13243708 were conditionally independent eQTLs in esophageal and breast tissue for *COPS6* and *ZKSCAN1,* respectively. However, we found no colocalization between the disease GWAS and eQTL signals for *COPS6, TRIM4,* or *ZKSCAN1* in the tissues of interest (PP4 < 0.312; **Table S8**). The lead eQTLs for *COPS6, TRIM4,* or *ZKSCAN1* were in weak with rs2897075 (r^2^ < 0.20).

For rs1105569, we tested 9 candidate protein-coding eGenes (**Table 2**) but did not find clear evidence of colocalization for any eGene in the tissues of interest, which was likely due to the extensive LD in the region (**Table S8**). Thus, out of five shared loci, baseline bulk eQTL data in the most relevant healthy tissue (i.e. lung) only revealed a clear causal gene (*MUC5B*) for rs35705950 (**Figure 3A-B**). Our focus on the shared genetic variants between critically ill COVID-19 and IPF prompted us to broaden our search to other relevant tissues, revealing that expression of *ATP11A* in whole blood and *DPP9* in fibroblasts may be relevant for the effect of rs12585036 and rs12610495, respectively, during pathogenesis (**Figure 3G**). Our results highlight a mystery in the field of human genetics – lack of colocalization between GWAS and eQTL signals.^8,53,54^ To address this, we next examined context-specific eQTL datasets.

### rs12585036 is an eQTL for ATP11A in lung tissue from a cohort of primarily ex- and current smokers

The Lung eQTL Study utilized lung samples from 1,111 patients, and most patients (85%) were either ex-smokers or current smokers (compared to 68% of all tissue donors in GTEx).^55-57^ We were interested in the Lung eQTL Study due to its larger sample size compared to GTEx and because smoking is a risk factor for ILDs including IPF.^58-60^ We hypothesized that this dataset might elucidate additional eQTL signals that were not detected in GTEx and better colocalize with the IPF and COVID-19 GWAS signals. Indeed, we found that a lead eQTL for *ATP11A* in the Lung eQTL Study, rs7998551 (p=7.68 x 10^-8^), was in strong LD with rs12585036 (r^2^ = 0.96) (**Figure 4A)**. We subsequently found evidence of colocalization between the disease GWAS and the *ATP11A*-eQTL signals (PP4 = 0.996 with IPF GWAS; 0.995 with COVID-19 GWAS; **Figure 4B-C; Table S9**) but lack of colocalization between the *ATP11A-*eQTL signals from the Lung eQTL Study and GTEx lung (PP4 = 0.344; **Table S9**). This demonstrated that lungs exposed to a particular environmental stressor, such as smoking, can reveal eQTLs and causal genes that would otherwise be overlooked. The C allele of rs12585036 was associated with higher *ATP11A* expression and was the risk allele in IPF but the protective allele for critically ill COVID-19. Thus, *ATP11A* appeared to have opposite effects on the pathophysiology of these two diseases.

**Figure 4:**
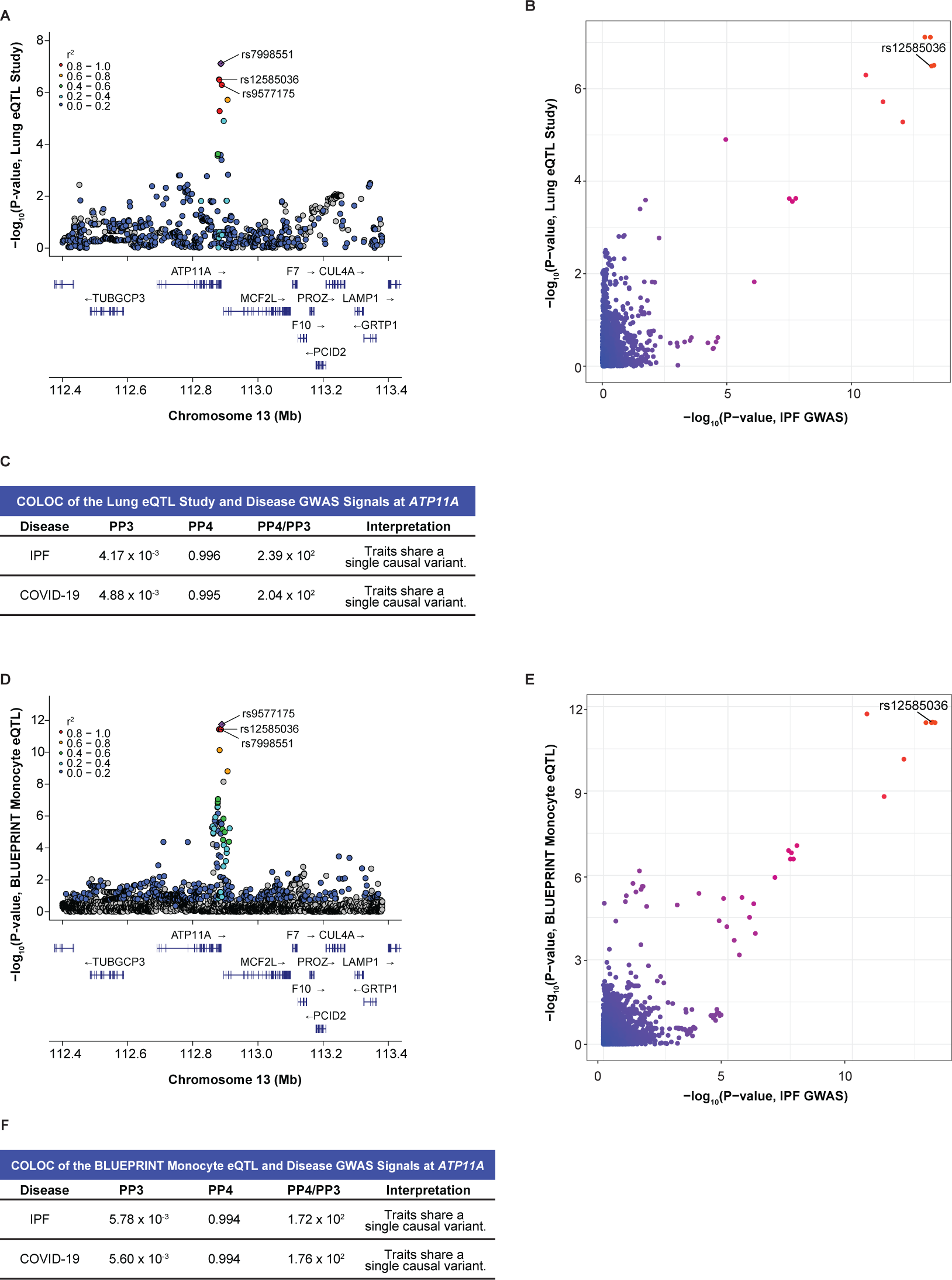
Colocalization at rs12585036 in a proinflammatory context and cell-type specific manner reveals *ATP11A* as a causal gene. (A) LocusZoom plot shows that rs12585036 is a top eQTL and in strong LD with the lead eQTL (rs7998551; purple diamond) for *ATP11A* in the Lung eQTL Study, which included primarily individuals with a smoking history. (B) Comparison of -log_10_(p-values) from the IPF GWAS and *ATP11A-*eQTLs in the Lung eQTL Study shows rs12585036 as a top shared SNP. (C) COLOC indicates strong colocalization between the disease GWAS and the Lung eQTL Study’s *ATP11A-*eQTL signals with PP4 > 0.900, and PP4/PP3 > 5.00. (D) rs12585036 is a top eQTL and in strong LD with the lead eQTL (rs9577175) for *ATP11A* in monocytes in the BLUEPRINT project. (E) Comparison of -log_10_(p-values) from the IPF GWAS and monocytic *ATP11A-*eQTLs shows rs12585036 as a top shared SNP. (F) COLOC indicates strong colocalization between the disease GWAS and monocytic *ATP11A-*eQTLs signals with PP4 > 0.900 and PP4/PP3 > 5.00. P-values are -log_10_ transformed. Linkage disequilibrium information for European populations was obtained from LDlink and is relative to the lead variant in each plot.

We found no colocalization at the *MUC5B* locus since the SNP array used in this study (Illumina Human1M-Duo BeadChip) did not include rs35705950, and there are no additional SNPs in LD with this SNP. We did not find colocalization at our other variants of interest.

### Monocytes are a critical cell type for the eQTL effect of rs12585036

The colocalization of rs12585036 with *ATP11A* expression in lung tissue primarily from smokers revealed a plausible environmental context where an eQTL effect is revealed. However, as these data were from bulk lung tissue, it was unclear what specific cell type drove this difference. Smoking causes inflammation and infiltration of immune cells, specifically monocytes and neutrophils, into the lungs.^61-63^ Therefore, we hypothesized the eQTL effect may be associated with a specific immune cell, which was also consistent with the modest colocalization we detected from bulk whole blood in GTEx (**Figure 3C-D**). The BLUEPRINT project^64^ allowed us to interrogate *ATP11A* eQTLs in monocytes, neutrophils, and CD4+ T cells. rs12585036 was an eQTL only in monocytes (p-value = 3.58 x 10^-12^ in monocytes vs. 1.38 x 10^-3^ in neutrophils and 3.89 x 10^-2^ in T cells; **Figure 4D**) and demonstrated strong colocalization with the disease GWAS (PP4 = 0.994; **Figure 4C-E; Table S9**). Directionality of the signals from the BLUEPRINT project and Lung eQTL Study were consistent with the C allele of rs12585036 being associated with higher *ATP11A* expression. Thus, our analyses used two context-specific eQTL datasets to reveal that *ATP11A* was the causal gene associated with the IPF and COVID-19 GWAS signals at rs12585036. Further, the eQTL signal appeared to be important specifically in monocytes, possibly within the proinflammatory context of smoking.

### rs12610495 is an eQTL for DPP9 in IPF lung

We sought to determine if there was colocalization with eQTLs specifically identified in lungs from individuals with IPF. We utilized the Borie et al. eQTL dataset which included lung samples from healthy controls (n = 188) and individuals with IPF who also reported a more extensive smoking history (n = 234).^28^ Borie et al. previously demonstrated colocalization of the eQTL, mQTL, and GWAS signals at rs35705950 within the *MUC5B* locus in both controls and IPF cases.^28^

We did not find evidence of colocalization between disease GWAS and control/IPF eQTL signals at rs12585036, rs2897075, and rs1105569. However, we found colocalization at rs12610495 for *DPP9* expression in IPF cases. Although the eQTL signals were in similar genomic locations for both controls and IPF cases, fine mapping further revealed that the lead variants between the two groups were distinct signals (**Figure 5A-D**). The lead variant in controls, rs758510 (p-value = 2.91 x 10^-6^ from FastQTL following PEER normalization), and the lead variant in the IPF cases, rs12462642 (6.65 x 10^-7^), exhibited weak LD (r^2^ = 0.15). In contrast, rs12462642 and rs12610495 (the lead GWAS variant; 6.52 x 10^-5^) were in stronger LD (r^2^ = 0.68). COLOC demonstrated moderately strong colocalization between the disease GWAS and IPF cases *DPP9-*eQTL signals (PP4 = 0.874 with IPF GWAS; 0.873 with COVID-19 GWAS) and weaker colocalization between the GWAS and control eQTL signals (PP4 = 0.661; 0.640; **Figure 5E-F; Table S9**). Plotting *DPP9* expression by rs12610495 genotype confirmed a more robust association of this lead GWAS SNP with expression in IPF cases (slope = -0.212; p-value = 1.96 x 10^-2^ using linear regression of pre-PEER normalized values) compared to controls (slope = -0.074; p-value = 0.526) and was consistent with the G allele being associated with lower *DPP9* expression (**Figure 5G-H**). Thus, two independent context-specific eQTL datasets, from fibroblasts and IPF lung, pointed to *DPP9* as the causal gene underlying rs12610495. These discoveries could be related as IPF lungs are known to have high levels of activated, and potentially abnormal, fibroblasts.^65-68^

**Figure 5:**
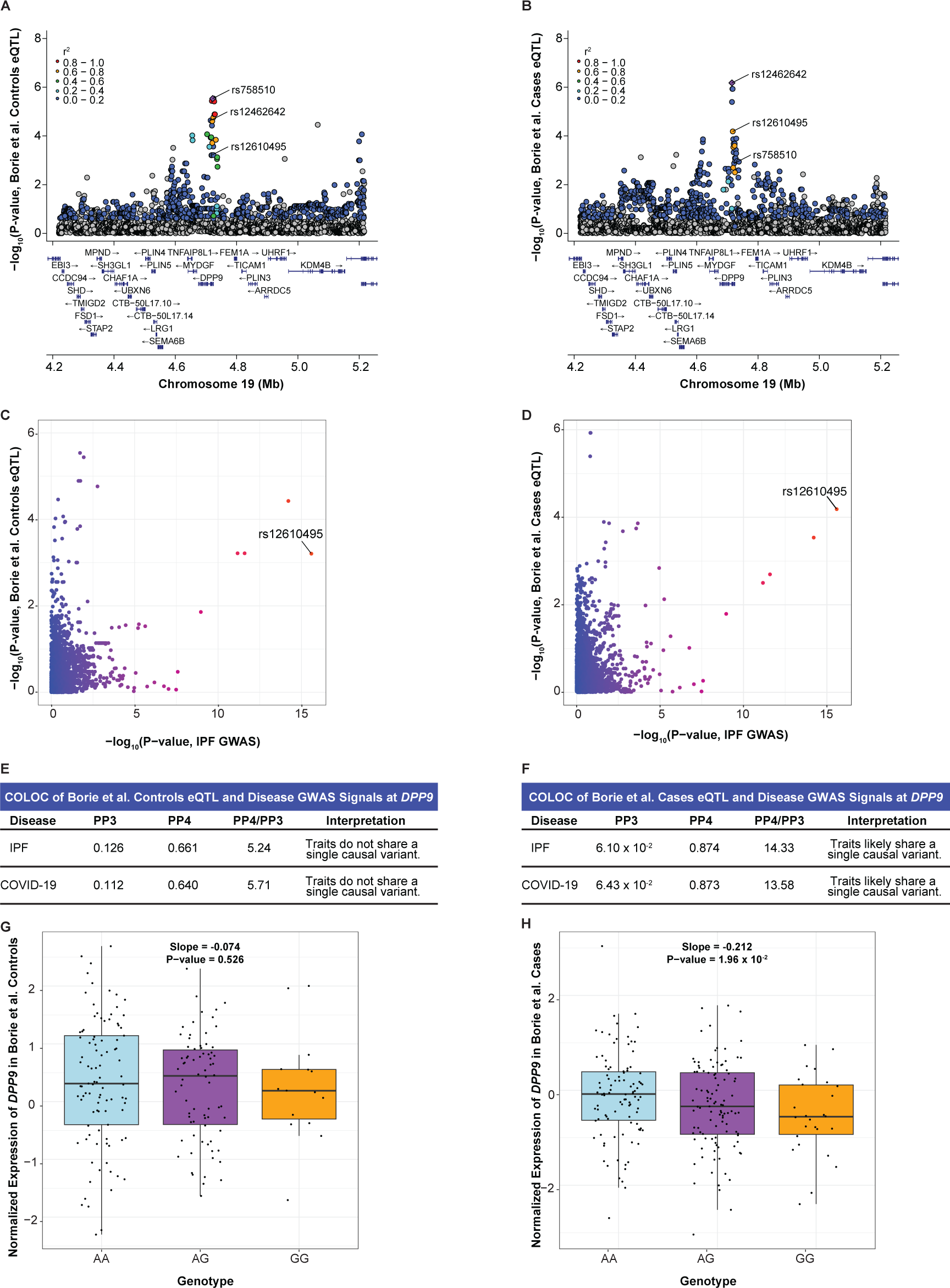
Colocalization at rs12610495 in IPF cases reveals *DPP9* as a causal gene. (A) LocusZoom plot shows that rs12610495 is not the lead eQTL nor in LD with the lead eQTL (rs758510; purple diamond) for *DPP9* in controls in the Borie et al. dataset. (B) rs12610495 is a top eQTL and in LD with the lead eQTL (rs12462642) for *DPP9* in IPF cases. (C) Comparison of -log_10_(p-values) from the IPF GWAS and *DPP9-*eQTLs in controls shows rs12610495 as a top shared SNP. (D) Comparison of -log_10_(p-values) from the IPF GWAS and *DPP9-*eQTLs in IPF cases shows rs12610495 as the lead shared SNP. (E) COLOC does not support colocalization between the disease GWAS and *DPP9-*eQTL signals in controls with PP4 < 0.700. (F) COLOC indicates colocalization between the disease GWAS and *DPP9-*eQTL signals in IPF cases with PP4 > 0.700 and PP4/PP3 > 5.00. (G) Box plot for *DPP9* expression in controls by genotype. (H) *DPP9* expression in IPF cases by genotype. P-values in LocusZoom plots are -log_10_ transformed. Linkage disequilibrium information for European populations was obtained from LDlink and is relative to the top variant in each plot. Slopes and p-values for genotype expression boxplots were obtained through linear regression modeling.

In summary, we used context-specific eQTL datasets of fibroblasts, monocytes, and lungs affected by smoking and IPF to show that *ATP11A* and *DPP9* were likely causal genes in critically ill COVID-19 and IPF. These results also revealed that *ATP11A* and *DPP9* were likely important in monocytes and fibroblasts, respectively. We also examined recent sc-eQTL datasets from Aquino et al. for COVID-19^69^ and Natri et al. for IPF^70^ but found no further evidence of colocalization, which could be due to low power (n = 80 Western European individuals in Aquino et al.; n = 48 controls, 66 IPF cases in Natri et al.).

### rs12610495 is an mQTL affecting methylation of the DPP9 promoter in IPF lungs

The stronger colocalization of GWAS and eQTL signals at rs12610495 in IPF cases suggested that there may be underlying gene regulatory differences between healthy and IPF lungs. Differences in DNA methylation can influence gene expression, and mQTL may reveal locations in the genome where patterns of DNA methylation are regulated by genetic variation.^71-73^ We utilized the Borie et al. mQTL dataset which included lung samples from healthy controls (n = 202) and IPF cases (n = 345). We found that rs12610495 was not in strong LD with rs10420225 (r^2^ = 0.25), the lead mQTL for cg07317664 in controls, but was in strong LD with rs2277732 (r^2^ = 0.95), the lead mQTL for cg07317664 in cases (**Figure 6A-B**). We did not observe colocalization between the disease GWAS and control cg07317664-mQTL signals (PP4 = 1.33 x 10^-2^ with IPF GWAS; 9.47 x 10^-3^ with COVID-19 GWAS) (**Figure 6C, 6E**). Like the eQTL colocalization at rs12610495, we found strong colocalization between the disease GWAS and IPF cases cg07317664-mQTL signals (PP4 = 0.980 with IPF GWAS; 0.763 with COVID-19 GWAS; **Figure 6D, 6F**). Plotting methylation of cg07317664 by rs12610495 genotype indicated a more robust association in IPF cases (slope = 1.9 x 10^-2^; p-value = 3.20 x 10^-9^ by linear regression using pre-PEER methylation values) compared to controls (slope = 1.2 x 10^-2^; p-value = 6.99 x 10^-4^) (**Figure 6G-H**).

**Figure 6:**
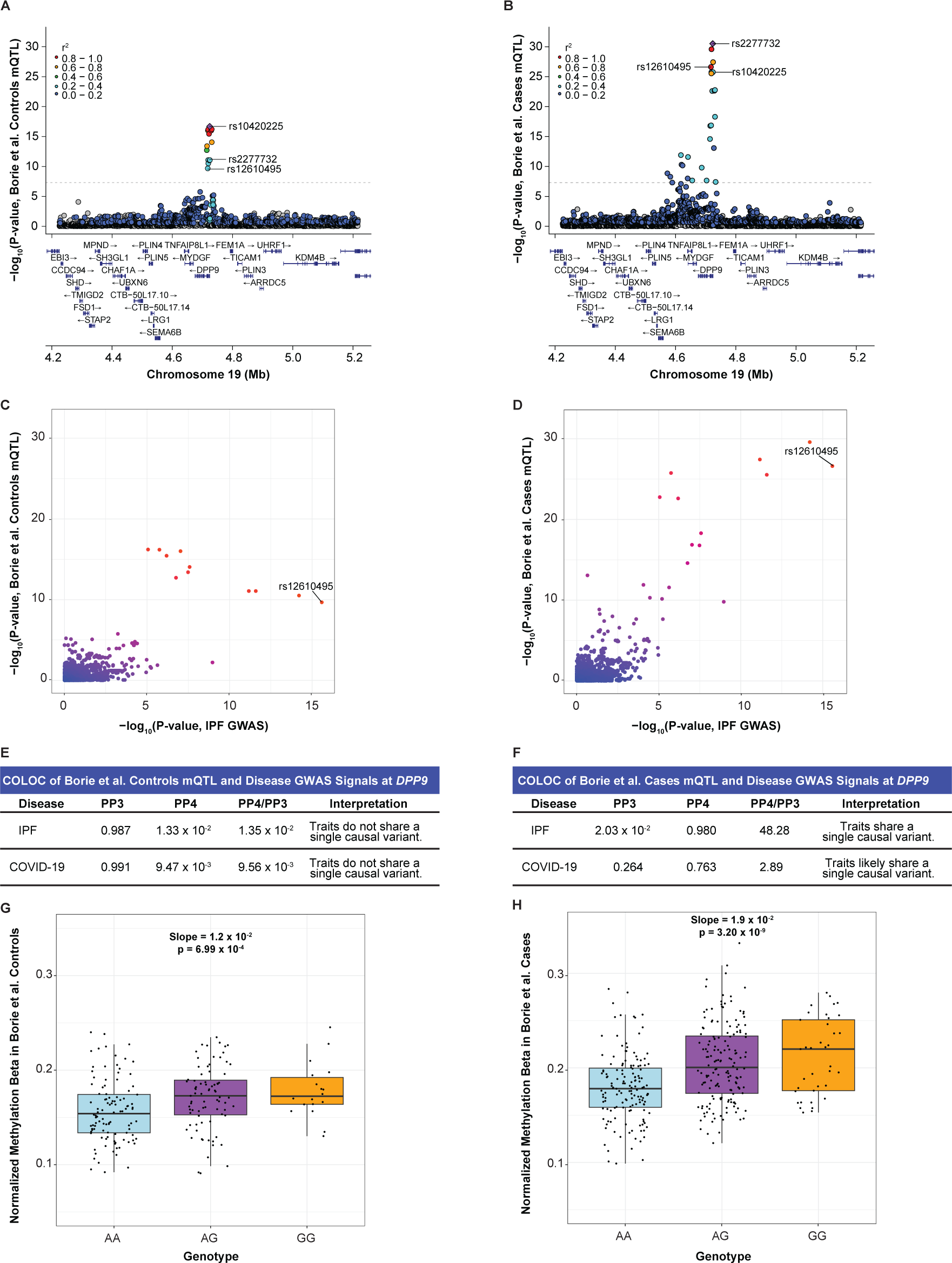
Colocalization at rs12610495 reveals a differentially methylated site (cg07317664) near the *DPP9* transcription start site in IPF cases. (A) LocusZoom plot shows that rs12610495 is not the lead mQTL nor in LD with the lead mQTL (rs10420225; purple diamond) for cg07317664 in controls in the Borie et al. dataset. (B) rs12610495 is a top mQTL and in LD with the lead mQTL (rs2277732) for cg07317664 in IPF cases. (C) Comparison of -log_10_(p-values) from the IPF GWAS and cg07317664-mQTLs in controls shows rs12610495 as a top shared SNP. (D) Comparison of -log_10_(p-values) from the IPF GWAS and cg07317664-mQTLs in IPF cases shows rs12610495 as the lead shared SNP. (E) COLOC does not support colocalization between the disease GWAS and cg07317664-mQTL signals in controls with PP4 < 0.700. (F) COLOC indicates colocalization between the disease GWAS and cg07317664-mQTL signals in cases with PP4 > 0.700. (G) Box plot for normalized methylation beta in controls by genotype. (H) Normalized methylation beta in IPF cases by genotype. P-values in LocusZoom plots are -log_10_ transformed. Linkage disequilibrium information for European populations was obtained from LDlink and is relative to the top variant in each plot. Slopes and p-values for genotype methylation beta boxplots were obtained through linear regression modeling.

Cg07317664 is 7319 base pairs upstream of the transcription start site of *DPP9,* leading us to hypothesize that methylation at this location may regulate *DPP9* gene expression. This hypothesis was consistent with the genetic and eQTL data as the G allele of rs12610495 was associated with greater methylation at a regulatory site of *DPP9* and decreased expression of *DPP9*, while the A allele was associated with less methylation and increased expression. Comparing the mQTL and eQTL signals in controls and cases separately demonstrated that colocalization occurred only in cases (PP4 = 0.404 in controls; 0.730 in cases; **Figure 7A-C**;). Further, we found that IPF cases, regardless of rs12610495 genotype, had significantly higher methylation levels at cg07317664 (slope = 0.0304, p-value < 2.2 x 10^-16^) and significantly lower expression of *DPP9* (slope = -0.561, p-value = 9.5 x 10^-9^; **Fig 7D-E)**, suggesting that genotype was just one factor contributing to reduced expression in IPF and underscoring the importance of considering epigenetic regulation.

**Figure 7:**
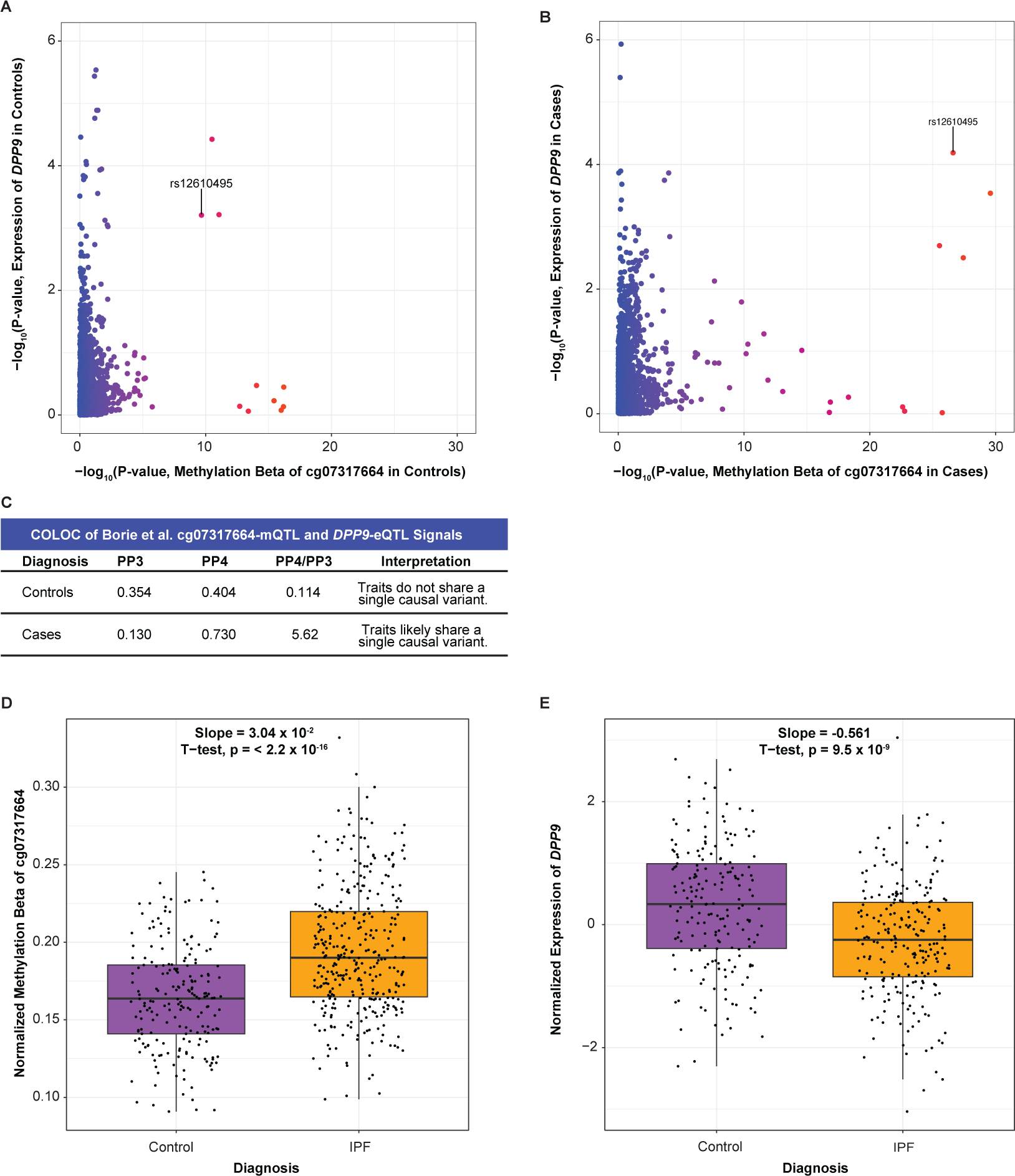
Colocalization at rs12610495 confirms the association between the cg07317664-mQTL and *DPP9*-eQTL signals in IPF cases. (A) Comparison of -log_10_(p-values) of cg07317664-mQTLs and *DPP9-*eQTLs in controls in the Borie et al. datasets shows that rs12610495 in not the lead SNP for either dataset. (B) Comparison of -log_10_(p-values) in IPF cases shows rs12610495 as a top shared SNP. (C) COLOC does not support colocalization between cg07317664-mQTL and *DPP9*-eQTL signals in controls with PP4 < 0.700 but supports colocalization in IPF cases with PP4 > 0.700. (D) Box plot for *DPP9* expression in lung by diagnosis. (E) Box plot for normalized methylation beta in lung by diagnosis.

### Causal genes in bulk diseased tissue

The putative causal genes we identified also demonstrated altered expression in IPF and COVID-19. Consistent with the genetic associations and as previously reported, we found that *MUC5B* expression was significantly increased in lung tissue from individuals with IPF when compared to tissue from controls in two independent studies (GSE213001 and GSE34692) (**Figure 8A-B**). In contrast, *ATP11A* and *DPP9* expression were significantly decreased in IPF lung tissue when compared to control tissue (**Figure 8A-B)**. Lower *DPP9* expression in IPF lung tissue was concordant with the disease GWAS and eQTL association as the G allele of rs12610495 was associated with lower *DPP9* and increased risk of IPF. In contrast, the directionality of the observation for *ATP11A* was discordant with the GWAS and eQTL association, which suggested the C allele of rs12585036 was associated with higher *ATP11A* and increased risk of IPF. However, two reasons may reconcile this observation for *ATP11A*. Firstly, *ATP11A* may be higher during the onset of disease and decrease in later stages, which was when most samples in these studies were collected. Later changes in *ATP11A* levels may reflect an effect of the disease on gene expression. Secondly, a single cell type may be underlying the genetic signal which is confounded when looking at bulk tissue and may also be depleted during later stages of disease.

**Figure 8:**
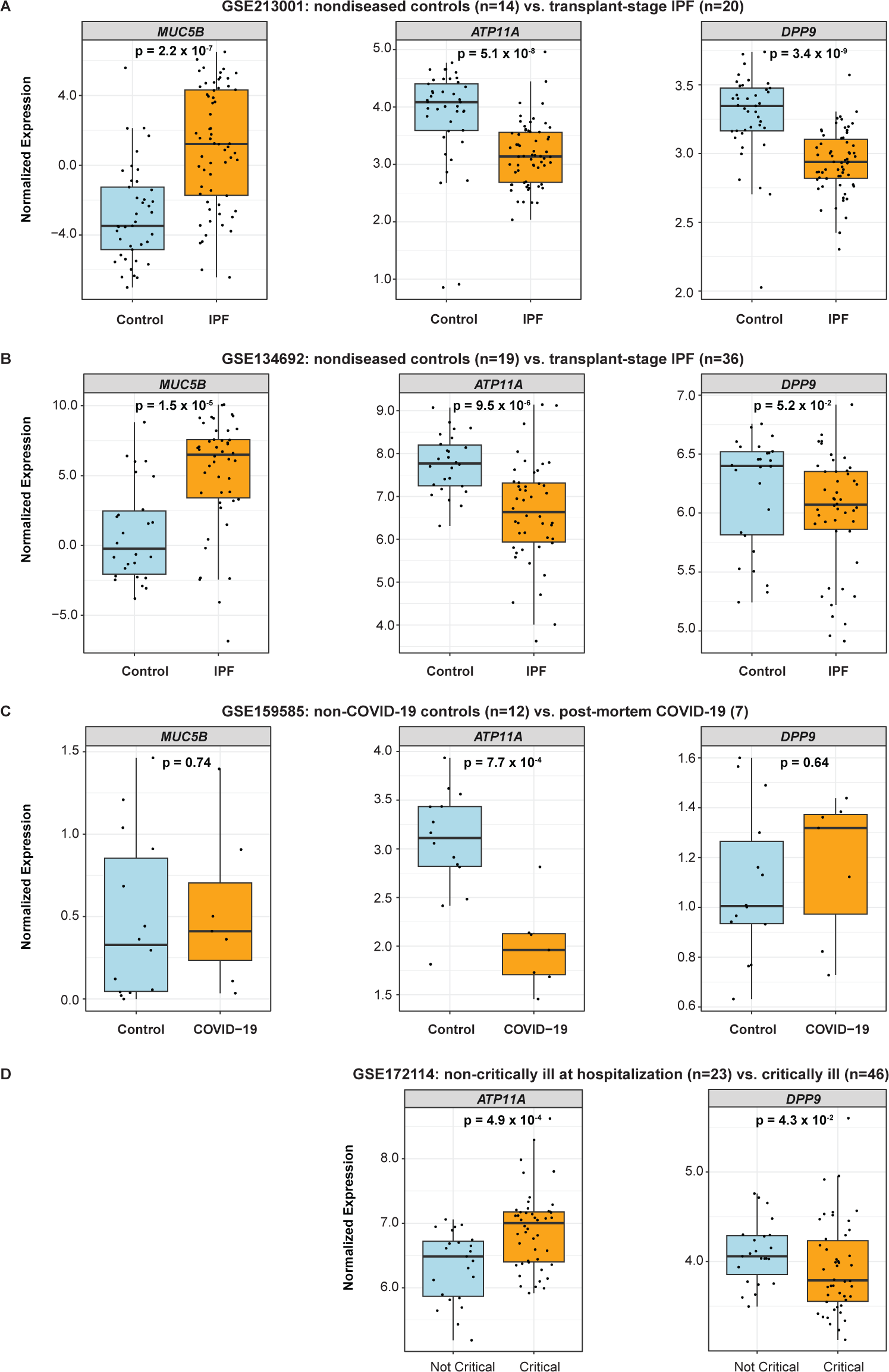
Assessment of causal genes in bulk control and diseased tissue from independent studies. Box plots depict normalized expression of causal genes by diagnosis from: (A) GSE213001, which included lung tissue from non-diseased controls (n=14) and IPF patients undergoing lung transplant (n=20). (B) GSE134692, which included lung tissue from non-diseased controls (n=19) and IPF patients undergoing lung transplant (n=36). (C) GSE159585, which included lung tissue from non-diseased controls (n=12) and from patients who succumbed to COVID-19 (n=7). (D) GSE172114, which included whole blood samples from hospitalized patients with non-critically ill COVID-19 (n=23) and patients in the intensive care unit with critically ill COVID-19 (n=46). P-values were calculated using the Mann-Whitney U test.

We did not observe significant differences in *MUC5B* or *DPP9* expression in lungs from patients who succumbed to COVID-19 when compared to control lungs, which could be due to the limited number of samples in the study we examined (GSE159585; **Figure 8C**). We found significantly lower *ATP11A* expression in COVID-19 lungs when compared to control lungs which was concordant with the GWAS and eQTL association as the T allele of rs12585036 was associated with low *ATP11A* and increased risk of COVID-19 (**Figure 8C**). However, in another study which measured gene expression in whole blood (GSE172114; **Figure 8D**), we found that *ATP11A* expression was higher in patients diagnosed with critically ill COVID-19 when compared to those with non-critically ill COVID-19, which could be due to an increase in myeloid lineage cells. As observed in the other datasets, *DPP9* is significantly lower in the blood of critically ill patients compared to the non-critically ill.

Altogether, these findings support the relevance of *ATP11A* and *DPP9* in IPF and COVID-19 pathogenesis, but directionality of how associated SNPs impact gene expression may differ in bulk diseased tissues possibly due to the impact of reverse causation of disease on gene expression, changes in cell type proportions, and/or the lack of temporal and spatial resolution. We therefore examined scRNA-Seq datasets.

### Causal genes in single cell types from diseased tissue

Using a scRNA-Seq dataset for IPF^38^ and snRNA-Seq dataset for lethal COVID-19^39^, we compared where the identified causal genes were expressed in healthy and diseased lung tissue. As differences in the expression of our identified causal genes could be due to either changes in cell type number or expression levels within a cell type, we first determined how cellular proportions differed between healthy and IPF or COVID-19 lungs. IPF lungs displayed decreases in type 1 and type 2 alveolar pneumocytes (AT1s and AT2s), and increases in other epithelial cell types (basal, ciliated, club, and goblet cells), myofibroblasts, peribronchial vascular endothelial cells, consistent with fibrosis and reduced healthy lung tissue (**Table S4)**. COVID-19 lungs displayed a significant increase only in macrophages but also notable increases in monocytes and fibroblasts, and decreases in AT1s, AT2s, and other airway epithelial cells (**Table S5**).

We subsequently determined what cell types expressed our causal genes. As expected, *MUC5B* was highly expressed in goblet cells and modestly in AT2s, other airway epithelial cells, and macrophages (**Table S6-7**). *ATP11A* was notably expressed in AT1s, AT2s, and macrophages and modestly expressed in fibroblasts, endothelial cells, monocytes, and other airway epithelial cells (**Table S6-7**). The expression in macrophages and monocytes was particularly intriguing given the colocalization of monocyte eQTL and GWAS signals at the *ATP11A* locus. *DPP9* was notably expressed in AT1s, AT2s, macrophages, and fibroblasts (**Table S6-7**).

Next, we examined differences in causal gene expression during IPF and COVID-19 in single cell types. Concordant with the GWAS and eQTL directionality, we found that *MUC5B* was significantly increased during IPF in goblet cells, AT2s, other airway epithelial cells, and macrophages (**Figure 9A**). In COVID-19, we found decreased, although statistically nonsignificant, *MUC5B* in airway epithelial cells, which includes goblet cells (**Figure 9B**), consistent with our genetic associations having opposite effects in these two diseases.

**Figure 9:**
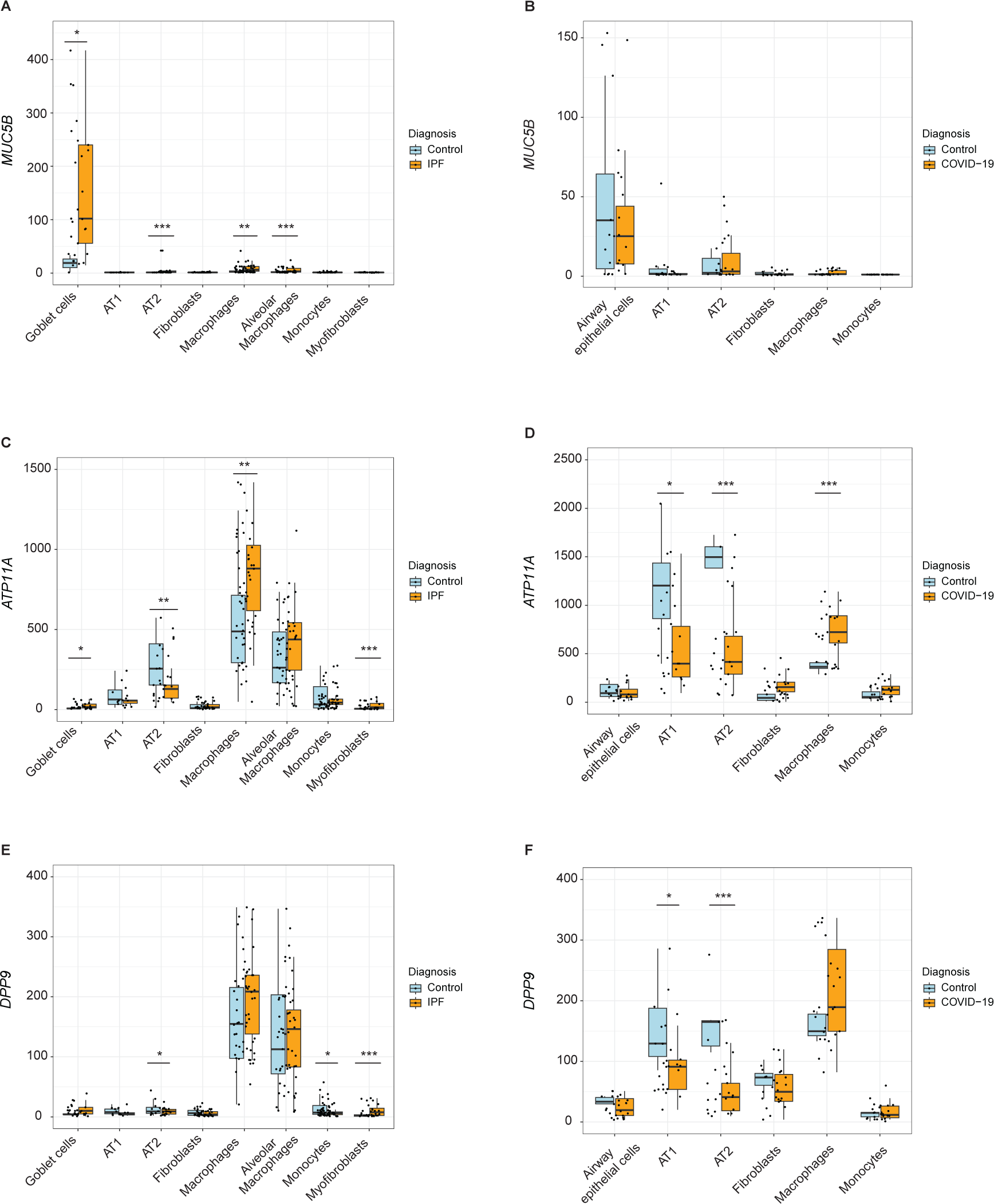
Assessment of causal genes at the single-cell resolution in control and diseased tissue. Box plots depict normalized expression of (A) *MUC5B*, (C) *ATP11A*, and (E) *DPP9* in an IPF single-cell RNA sequencing study by Adams et al. which included controls (n=28) and IPF patients (n=32). Box plots depict normalized expression of (B) *MUC5B*, (D) *ATP11A*, and (F) *DPP9* in a lethal COVID-19 single-nucleus RNA sequencing study by Melms et al. which included controls (n=7) and COVID-19 patients (n=19). P-values were calculated using Wald test with P values adjusted for multiple comparisons using the Benjamini-Hochberg method (*Padj < .05; **Padj < .001; ***Padj < .0001).

We hypothesized that *ATP11A* would be increased in epithelial cells and/or myeloid lineage cells based on the GWAS and eQTL colocalization in lung tissue primarily from smokers and in monocytes. However, we found significantly lower expression in AT2s (**Figure 9C; Table S6**), which may be due to the depletion of epithelial cells during IPF. Consistent with our hypothesis, *ATP11A* was significantly increased during IPF in macrophages, a large proportion of which are derived from monocytes recruited to the lung during IPF,^74^ as well as in other airway epithelial cells and myofibroblasts, which are a key driver of fibrosis (**Figure 9C; Table S6**).^65^ During COVID-19, *ATP11A* was significantly decreased in AT1s, AT2s, other airway epithelial cells, concordant with the GWAS and eQTL data (**Figure 9D; Table S7)**.

We hypothesized that *DPP9* would be decreased during IPF and COVID-19 in fibroblasts and other cell types that increase in proportion during IPF based on our GWAS and eQTL colocalization in fibroblasts and IPF lung. *DPP9* was significantly decreased in AT2s and monocytes during IPF (**Figure 9E; Table S6**). However, we found that *DPP9* was significantly increased in myofibroblast during IPF, which may be due to the hyperproliferation of this cell type during IPF (**Figure 9E; Table S6)**. During COVID-19, we found that *DPP9* was significantly decreased in AT1s and AT2s (**Figure 9F; Table S7)**.

## DISCUSSION

GWAS provide a powerful approach to gather human genetic evidence to increase success in drug development, but such efforts require the identification of the underlying causal genes.^1^ Molecular QTL studies were designed to bridge the gap between GWAS variants and causal genes, revealing molecular effects on gene expression, splicing, and methylation. However, studies have found limited colocalization between eQTL and human disease GWAS signals. This has been speculated to be due to multiple factors including lack of power in detecting GWAS and eQTL signals; inappropriate context with regards to cell type, developmental stage, and response; nonlinear relationships between eQTL effect and phenotype; overlap of eQTL signals due to multiple signals; and strong LD between variants limiting deconvolution of causality (e.g. rs1105569 within the 17q21.31 inversion supergene). Here, we found evidence that consideration of cell type and disease can improve colocalization of GWAS and QTL signals to reveal causal genes and where they may function during pathogenesis. Whereas eQTLs from healthy lung revealed *MUC5B* as the causal gene underlying the association of rs35705950 with critically ill COVID-19 and IPF, *MUC5B* may be atypical in that its expression is largely restricted to mucus-secreting cells and the effect size of the eQTL is large,^28^ making accurate mapping of this eQTL feasible with smaller numbers and in bulk tissue. Examination of eQTL data from isolated cell types and lung disease states was necessary to reveal *DPP9* and *ATP11A* as two additional causal genes. Thus, well-powered tissue- and disease-specific eQTL datasets will continue to reveal additional causal genes for human diseases.

For each of the five shared loci between critically ill COVID-19 and IPF, identification of the likely causal gene leads to testable hypotheses for how each gene impacts these two diseases. This has been demonstrated for the *MUC5B* locus in IPF. The variant rs35705950 is within an enhancer that is differentially regulated and binds the transcription factor FOXA2 to regulate *MUC5B* expression.^52^ Additional colocalization studies at rs35705950 between IPF GWAS, control and IPF eQTL, and mQTL signals suggested that higher methylation at a repressor region further contributes to increased *MUC5B* expression in cases of IPF.^28^ *In vivo* studies have functionally demonstrated that excessive MUC5B production impairs mucociliary clearance and increases the fibroproliferative response, while reducing mucin production may ameliorate fibrosis.^75^ Though increased mucus plugging was observed in COVID-19 lung autopsies,^76^ the same allele associated with increased *MUC5B* is protective against severe COVID-19, suggesting increased mucus production in this acute context is protective. The opposite risk alleles of *MUC5B* (and *ATP11A*) for IPF and critically ill COVID-19 also underscores that though these two diseases share five of the same genetic determinants, the underlying pathophysiology is unique—critically ill COVID-19 and PCPF is not simply IPF due to a known etiology. For the remaining four loci overlapping between COVID-19 and IPF, identification of causal genes has now begun to similarly elucidate mechanisms of diseases.

The causal gene associated with rs2897075 has been difficult to identify due to lack of colocalization between GWAS and eQTL signals. We speculate that the most likely causal genes are *TRIM4* and *ZKSCAN1.* rs2897075 is an eQTL for *TRIM4* in several tissues, including the lung, and decreased expression is associated with the risk allele of critically ill COVID-19 and IPF. TRIM4 has been reported to polyubiquitinate viral pattern recognition receptors including RIG-I and MDA5 and promoting type I interferon production during Sendai virus^77^ and SARS-CoV-2^78^ infections, respectively. rs2897075 is also an eQTL for *ZKSCAN1* in fibroblasts, and decreased expression is associated with the risk allele of IPF and critically ill COVID-19. Interestingly, *ZKSCAN1* mRNA can be backspliced into a circular RNA (circRNA), a type of noncoding RNA that can act as sponges for microRNA and proteins and may contribute to fibrosis.^79,80^ Whether rs2897075 is associated with levels of *ZKSCAN1* circRNA is unknown, though both *ZKSCAN1* and *circZKSCAN1* have been implicated in liver fibrosis.^81^ Although several studies have reported rs2897075 as a genome-wide significant hit in IPF GWAS, its mechanism and phenotypic effect has not been reported to our knowledge. Importantly, we noted that although rs2897075 was not an eQTL in the Borie et al. eQTL dataset, a variant in strong LD rs6963345 (r^2^ = 1.00) was associated with numerous mQTLs, suggesting that this SNP may influence DNA methylation.^28^

The directionality of the rs12585036 alleles indicates that increased *ATP11A* expression is associated with increased risk of IPF while decreased expression is associated with increased risk of critically ill COVID-19. Our colocalization data suggest that the association at the *ATP11A* locus may be due to expression in monocytes and derived cells in the lung. Cycles of alveolar damage, recruitment of monocytes, and their differentiation into monocyte-derived alveolar macrophages occur repeatedly during IPF.^74^ Specifically, M2 macrophages promote fibrosis, possibly through secretion of TGF-beta^82^ and other profibrotic factors.^74^ ATP11A, or ATPase Phospholipid Transporting 11A, is a member of the P4-ATPase family of lipid flippases, responsible for translocating aminophospholipids on the cellular membrane.^83,84^ We propose two mechanisms that could explain ATP11A’s involvement in the pathogenesis of IPF and COVID-19. Firstly, ATP11A is cleaved during apoptosis and results in phosphatidylserine’s translocation to the outer leaflet of the cell membrane, which is an “eat me” signal during efferocytosis.^83-86^ Increased apoptosis of epithelial cells and resistance to apoptosis in myofibroblasts are hallmarks of IPF, correlating with aberrant collagen deposition.^87,88^ During IPF, we speculate that increased ATP11A may contribute to impaired efferocytosis, an observation reported in murine models^89^ and IPF patient samples^90^. Secondly, ATP11A has also been reported to promote internalization of toll-like receptor 4 (TLR4), which is highly expressed on both alveolar pneumocytes and myeloid lineage cells, preventing hypersecretion of proinflammatory cytokines after stimulation with lipopolysaccharide.^91^ This may skew macrophages toward profibrotic M2 polarization. Additionally, TLR4 signaling contributes to proper AT2 renewal and lung repair.^92^ In the case of COVID-19, the ectodomain of the SARS-CoV-2 spike protein has been consistently reported to stimulate TLR4.^93,94^ Without adequate internalization of TLR4 by ATP11A, downstream immune signaling may be overactivated, leading to hypersecretion of proinflammatory cytokines. Thus, we propose that ATP11A may affect IPF and COVID-19 by functioning in both epithelial and myeloid cells and eliciting impaired cell death and immune pathways (**Figure 10A**). Additionally, the opposite effects of rs12585036 and rs35705950 during IPF and COVID-19 highlight the trade-offs of specific genetic variants in the face of different threats.

**Figure 10:**
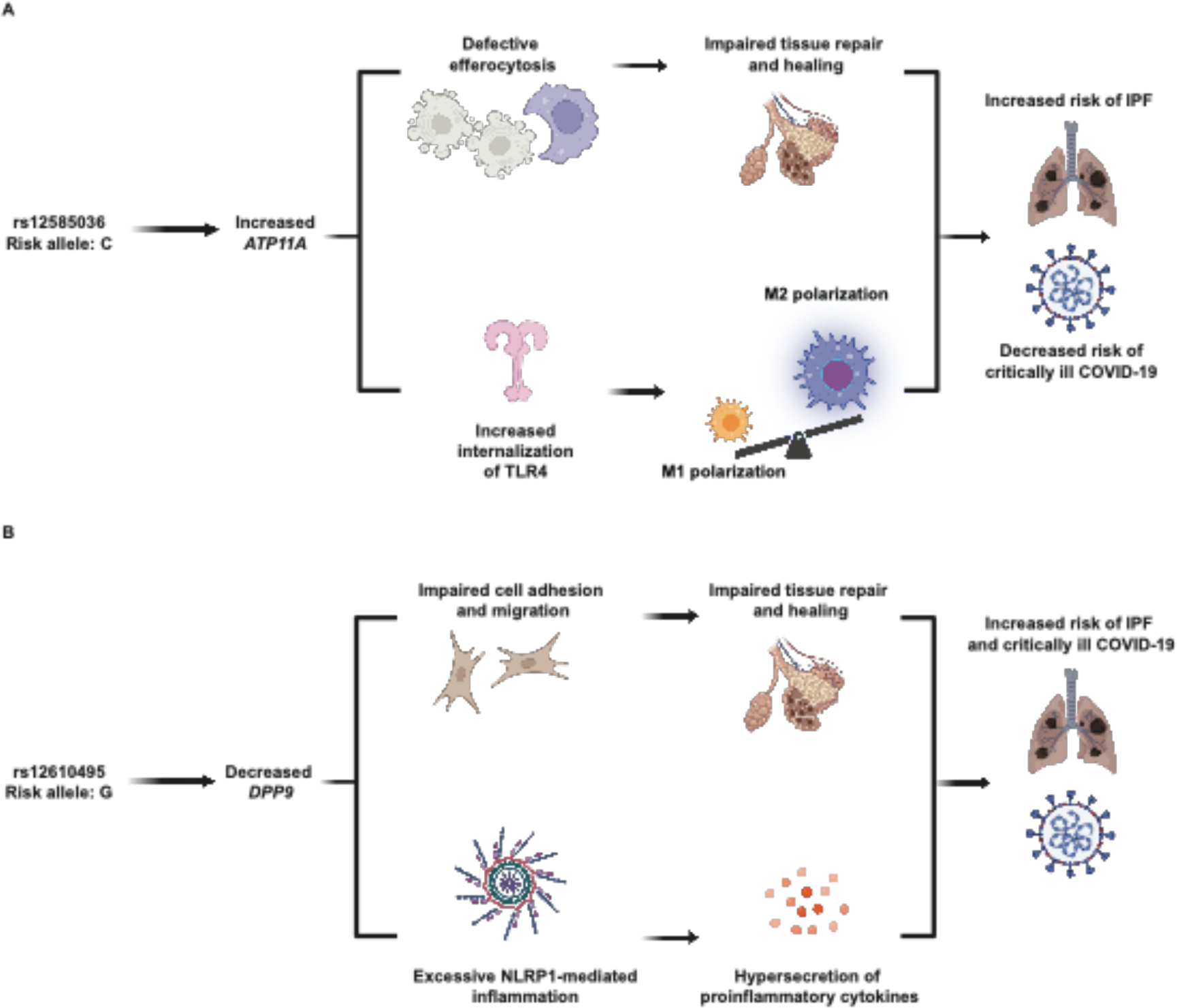
Hypothesized mechanisms linking disease-associated SNPs to causal genes, *ATP11A* and *DPP9*. (A) The C allele of rs12585036 is associated with increased *ATP11A*, leading to 1) defective efferocytosis and impaired lung tissue repair and 2) increased internalization of toll-like receptor 4 and increased M2-macrophage polarization. These mechanisms lead to increased risk of IPF and protection from COVID-19. (B) The G allele of rs12610495 is associated with increased *DPP9*, leading to 1) impaired cell adhesion and migration affecting lung tissue repair, and 2) excessive NLRP1-inflammasome activity and proinflammatory cytokine production. These mechanisms lead to increased risk of both IPF and COVID-19.

Colocalization of disease GWAS and eQTL signals indicates that decreased *DPP9* expression is associated with increased risk of COVID-19 and IPF and that fibroblasts may drive the association at least in IPF lung. DPP9, Dipeptidyl Peptidase 9, is a serine peptidase that is in the same family as DPP4, which is well-known as a target of hypoglycemic drugs and as a receptor for the coronavirus causing Middle Eastern Respiratory Syndrome. DPP9 helps maintain the inactive conformation of the NLRP1 inflammasome, inhibiting activation and downstream pyroptosis.^95-97^ In addition to regulating inflammasome activity, DPP9 also regulates survival, proliferation, migration, and adhesion, including in dermal fibroblasts.^98^ Interestingly, fibroblast activating protein (FAP) belongs to the same serine protease subfamily as DPP9. However, unlike the other DPP proteins, FAP is largely expressed by cancerous cells, inducing EMT. In IPF, FAP is specifically localized to fibrotic foci and not in normal tissue.^99^ DPP9 interacts with FAP directly in oral squamous cell carcinoma, and *DPP9* overexpression can inhibit the EMT activity induced by FAP.^100^ We previously reported that based on transcriptomics from patients infected with COVID-19 that *DPP9* decreased concordantly with resolution of infection, and therefore, may be dampening the inflammatory response.^11^ We suspect that lower DPP9 may exacerbate pathogenesis of IPF and COVID-19 by preventing excess NLRP1-mediated inflammation and weakening cell-to-cell adhesion, promoting a dysregulated cycle of inflammation and fibrosis (**Figure 10B**).

While the evidence we have presented suggests that variants in *ATP11A* and *DPP9* impact COVID-19 and IPF pathogenesis by altering expression of these genes, recent evidence from Nakanishi et al. demonstrated that the sQTL data for these loci also colocalize with severe COVID-19.^101^ Future studies will be necessary to understand the effect of these risk variants on splicing, but we note that each sQTL is predicted to only impact a single non-canonical isoform based on Ensembl annotation. The isoform associated with *DPP9* (ENST00000599248) is not protein-coding; the associated isoform for *ATP11A* (ENST00000415301) is highly truncated. Thus, *ATP11A* and *DPP9* demonstrate a new “problem” in human genetics; the same human disease signal may colocalize with multiple molecular QTL signals, expanding the list of possible causal genes that underlie a GWAS signal. eQTL and sQTL data may also demonstrate two consequences of the same SNP as altering splicing can certainly impact overall gene expression.

Identifying shared genetic associations and their causal genes can reveal new insight into the pathogeneses of COVID-19 and IPF that could spur further therapeutic development. Indeed, MUC5B has been proposed as a target for IPF treatment since the genetic association was first reported.^102^ For the causal genes with new evidence presented here, *DPP9* may be particularly attractive, and inhibitors in development have been speculated to be therapeutic for pulmonary fibrosis.^103^ However, for treatment of COVID-19 and IPF, increasing *DPP9* expression or activity seems to be the desired outcome. It remains to be tested if recombinant DPP9 or increasing expression in fibroblasts is beneficial in critically ill COVID-19 or IPF.

## Data Availability

All data produced in the present study are available upon reasonable request to the authors

## ACKNOWLEDGEMENTS

TD, LW, AGJ, and DCK were supported by R01AI118903 and R01AI170089. TD was supported by a TriCEM Graduate Student Award and the Gertrude B. Elion Mentored Medical Student Research Award. We thank Dr. Richard J. Allen and colleagues for sharing the IPF meta-GWAS summary statistics.

